# Spatiotemporal relationships between extreme weather events and arbovirus transmission across Brazil

**DOI:** 10.1101/2025.08.01.25332773

**Authors:** Victoria M. Cox, Felipe Campos de Melo Iani, Wes Hinsley, Pedro S. Peixoto, Flavio Codeço Coelho, Carlos Augusto Prete, Megan O’Driscoll, Neil Ferguson, Samir Bhatt, Nuno R. Faria, Ilaria Dorigatti

## Abstract

Brazil experiences large-scale annual outbreaks of dengue, chikungunya, and Zika virus infection, which are transmitted to humans by *Aedes* mosquitoes. Dengue is expanding into the north and south of Brazil and incidence of infection has been increasing over the last decade. Whilst previous analyses at the state and microregion level demonstrated that climate and environmental conditions affect dengue transmission, summary statistics computed over these large geographical areas can remove significant effects and mask local drivers of arbovirus transmission, which are needed for operational decisions on disease surveillance and control at the municipality level.

We analysed the weekly case notification timeseries of chikungunya, Zika, and dengue virus reported at the municipality level for Brazil (n=5550 administration units) from 2013 to 2020 using spatiotemporal mixed-effects regression models. We used this highly granular data to test the association between arbovirus incidence and 139 variables capturing meteorological conditions, environment, El Niño Southern Oscillation (ENSO) patterns, human connectivity, and socioeconomics of the resident population. We assessed which factors best captured the historic transmission dynamics of chikungunya, Zika, and dengue including extremes of rainfall and temperatures at different time lags.

Our findings highlight the joint health impact of poverty and extreme weather conditions on arbovirus infections in Brazil. We found that reduced socioeconomic indicators such as household income and access to adequate sanitation were associated with increased arbovirus incidence. Higher temperatures were positively associated with arbovirus incidence up to limiting negatively associated maxima. Summary statistics representing extreme conditions, such as the absolute maxima of environmental temperatures, ENSO anomalies, and long-term periods of extreme wetness or drought, were among the key predictors of arbovirus incidence.

The findings presented in this study shed new light on the long-term drivers of dengue transmission at unprecedented spatiotemporal resolution, which in future work can be used to reconstruct the attribution of anthropogenic climate change and to evaluate how climate change scenarios are expected to affect arbovirus dynamics going forward.

A version of this abstract in Portuguese can be found in the Supplementary Material.

## Introduction

The frequency and intensity of dengue (DENV), chikungunya (CHIKV), and Zika (ZIKV) virus epidemics is increasing globally ^1^. This is mainly driven by the geographical expansion of *Aedes* mosquitoes, which have long been established across Southeast Asia, South America, and Africa into temperate regions ^2–4^. These trends are expected to further increase under changing climate, urbanisation, and globalisation ^5–9^.

CHIKV was first detected in Brazil in August 2014 ^10^, and spread rapidly across the country ^11,12^. It is estimated that ZIKV was cryptically introduced in northeast Brazil in 2014 ^13,14^, ahead of the 2015-2018 epidemic ^14^. Compared to CHIKV and ZIKV which were introduced in Brazil within the last decade, DENV re-emerged in Brazil in 1981 and has since become hyperendemic ^15,16^, causing large-scale outbreaks each year, with around 2.1 million reported cases in 2019 alone ^17^. These arboviruses have recently been expanding into previously unaffected regions in Brazil: DENV is expanding in the north and south of Brazil ^18,19^ and it has been reported that indigenous populations living in the Amazon will be at higher risk of DENV ^20^. Between 2018 and 2022, 481 new municipalities experienced community-level DENV transmission ^19^ and between 2021 and 2023, 570 new municipalities reported CHIKV cases for the first time ^21^.

Meteorological conditions differ greatly between regions; for example, the North and Northeast experience warmer temperatures and smaller temperature ranges compared to the Centre-West, South, and Southwest. Moreover, the timing of arbovirus outbreaks is typically highly heterogeneous across Brazil, with states in the Centre-West region experiencing DENV transmission peaks earlier than those in the Northeast region ^22^. Spatial and temporal variations in transmission dynamics beyond seasonal trends have been recognised to be associated with urbanisation and meteorological conditions ^18,23^, in keeping with our understanding of mosquito biology. Mosquitoes are ectotherms whose life traits determining survival, fecundity, development, feeding, and transmission potential are sensitive to changes in environmental conditions, most notably temperature ^24–26^ but also humidity ^27,28^. For example, increases in temperature accelerate larval and pupal development rates and decrease extrinsic incubation periods in *Aedes* mosquitoes, however at threshold high temperatures, thermal stress and desiccation reduces adult survival rates ^27–29^. In Singapore, high levels of rainfall have been shown to increase suitable habitat for egg-laying and immature mosquitoes, up to an optimal level where excess rain flushes away breeding reservoirs ^30^. On the other hand, it has been hypothesised that drought may induce communities to store water in open containers where mosquitoes can breed ^31^. It is therefore important to account for heterogeneities in meteorological conditions including extreme events on arbovirus transmission, because analyses conducted using summary statistics of meteorological variables computed over large geographical units and/or time periods risk smoothing, confounding. or removing potentially significant effects, thus limiting our understanding of the impact that local conditions have on transmission and seasonal dynamics. In this work we used spatiotemporal regression models fitted to weekly historical CHIKV, DENV, and ZIKV case notification timeseries reported at the municipality level across Brazil from 2013 to 2020, to investigate the association between climate, environment, human mobility, socioeconomics, and arbovirus incidence at unprecedented spatial and temporal resolution.

## Results

We observed regular annual peaks of each of the three arboviruses at the subnational level, with consistently reduced DENV incidence in 2017-2018 following the 2015-2016 ZIKV outbreak (Figure 1B). The intra-annual patterns show that the relative incidence of reported cases peaks at the start of the year for most states (Figure 2), with flatter, less cyclic patterns for states in the North and Northeast regions. Across space, we observe clear clustering of arbovirus incidence, with municipalities in the Northeast reporting higher numbers of CHIKV cases and municipalities in the South reporting fewer DENV cases (Figure 1A).

**Figure 1.**
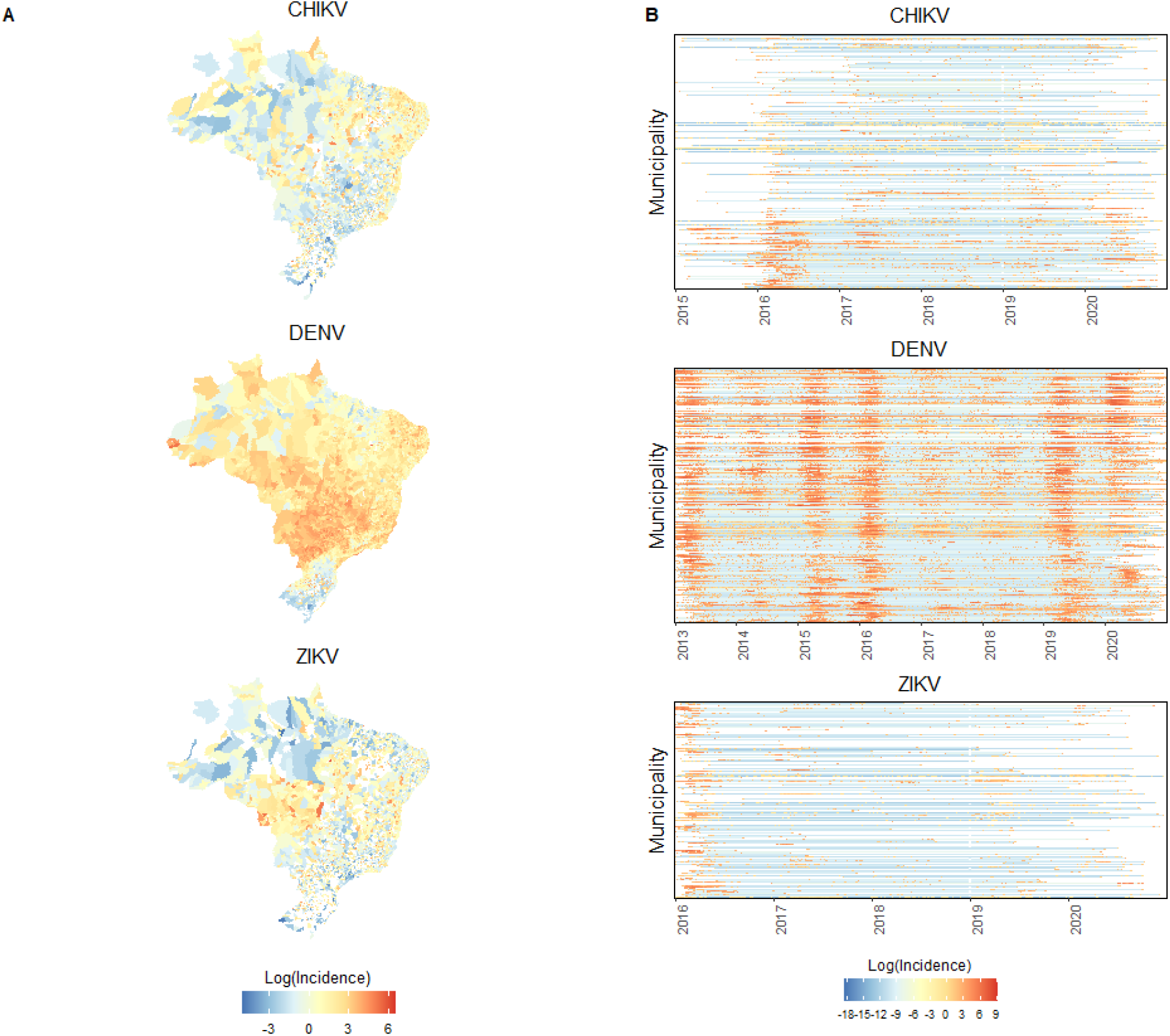
Summary of reported cases of chikungunya (CHIKV), dengue (DENV), and Zika (ZIKV) in Brazil between 2013 and 2020. (A) Incidence of reported cases (per 100,000 persons) in each municipality of Brazil, averaged over all time points, shown on the log scale. White indicates municipalities which did not report cases over the timeseries. (B) Weekly incidence of reported cases (per 100,000 persons) per municipality in Brazil, shown on the log scale. The municipalities are ordered on the y-axis by longitude from west at the top to east at the bottom.

**Figure 2.**
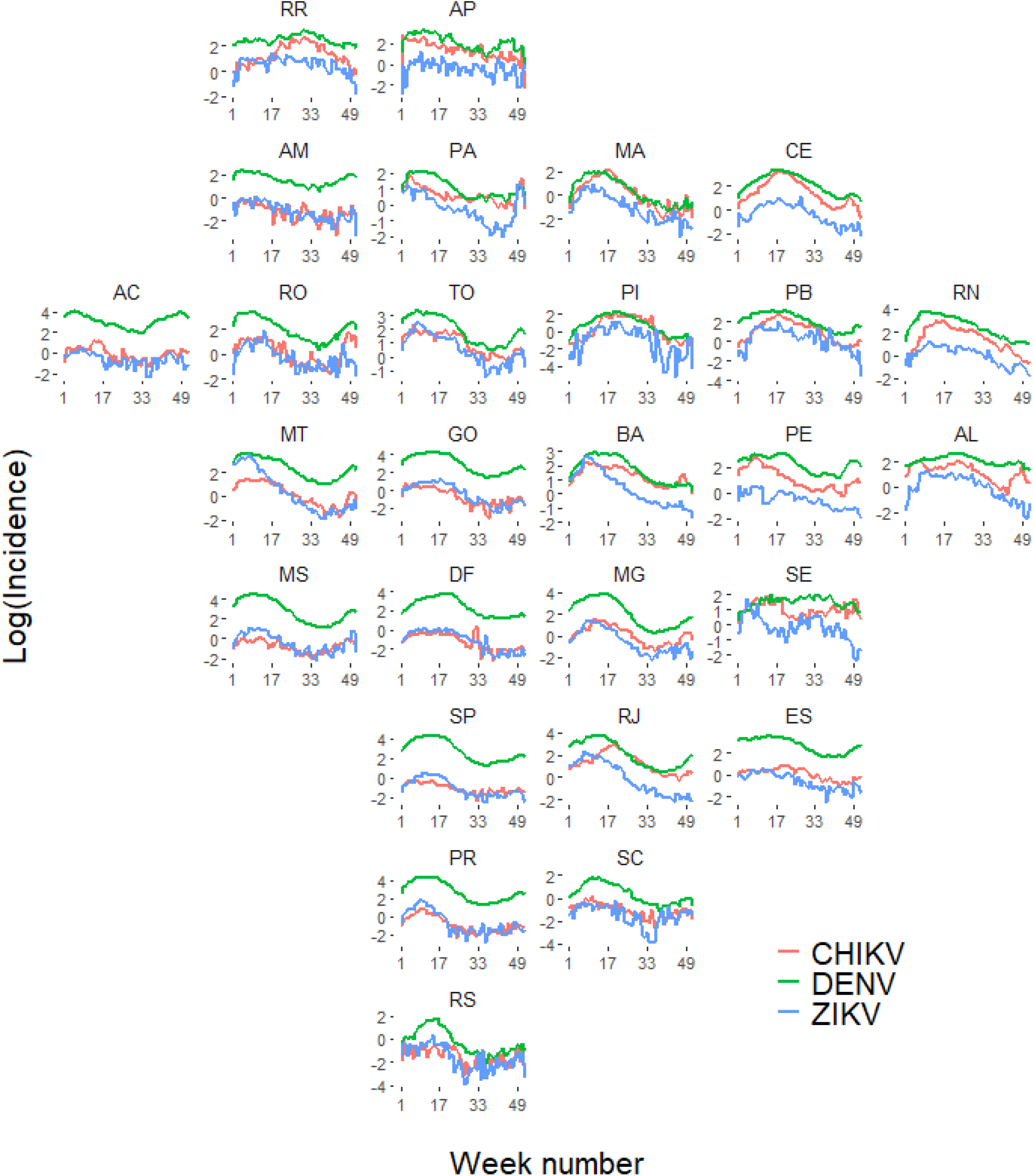
Summary of reported cases of chikungunya (CHIKV), dengue (DENV), and Zika (ZIKV) in Brazil between 2013 and 2020. Incidence of reported cases (per 100,000 persons) in each week of the year, averaged over the years and shown on the log scale. The panels represent states and are arranged geographically. Key: AC = Acre, AL = Alagoas, AM = Amazonas, AP = Amapá, BA = Bahia, CE = Ceará, DF = Distrito Federal, ES = Espírito Santo, GO = Goiás, MA = Maranhão, MG = Minas Gerais, MS = Mato Grosso do Sul, MT = Mato Grosso, PA = Pará, PB = Paraíba, PE = Pernambuco, PI = Piauí, PR = Paraná, RJ = Rio de Janeiro, RN = Rio Grande do Norte, RO = Rondônia, RR = Roraima, RS = Rio Grande do Sul, SC = Santa Catarina, SE = Sergipe, SP = São Paulo, TO = Tocantins.

The results obtained with the spatiotemporal regression models developed in this analysis reveal that sea surface temperature anomalies due to El Niño Southern Oscillation (ENSO) was the best predictor of CHIKV and ZIKV incidence and the second best for DENV incidence after the absolute maximum temperature (Tmax_max) lagged by 0-2 weeks (Tables 1 and S1). The associations between ENSO anomalies, which capture global deviations in climate patterns – between a cold phase (La Niña, where the Nino3.4 index is negative) and a warm phase (El Niño, where Nino3.4 is positive) – were positive.

**Table 1.**
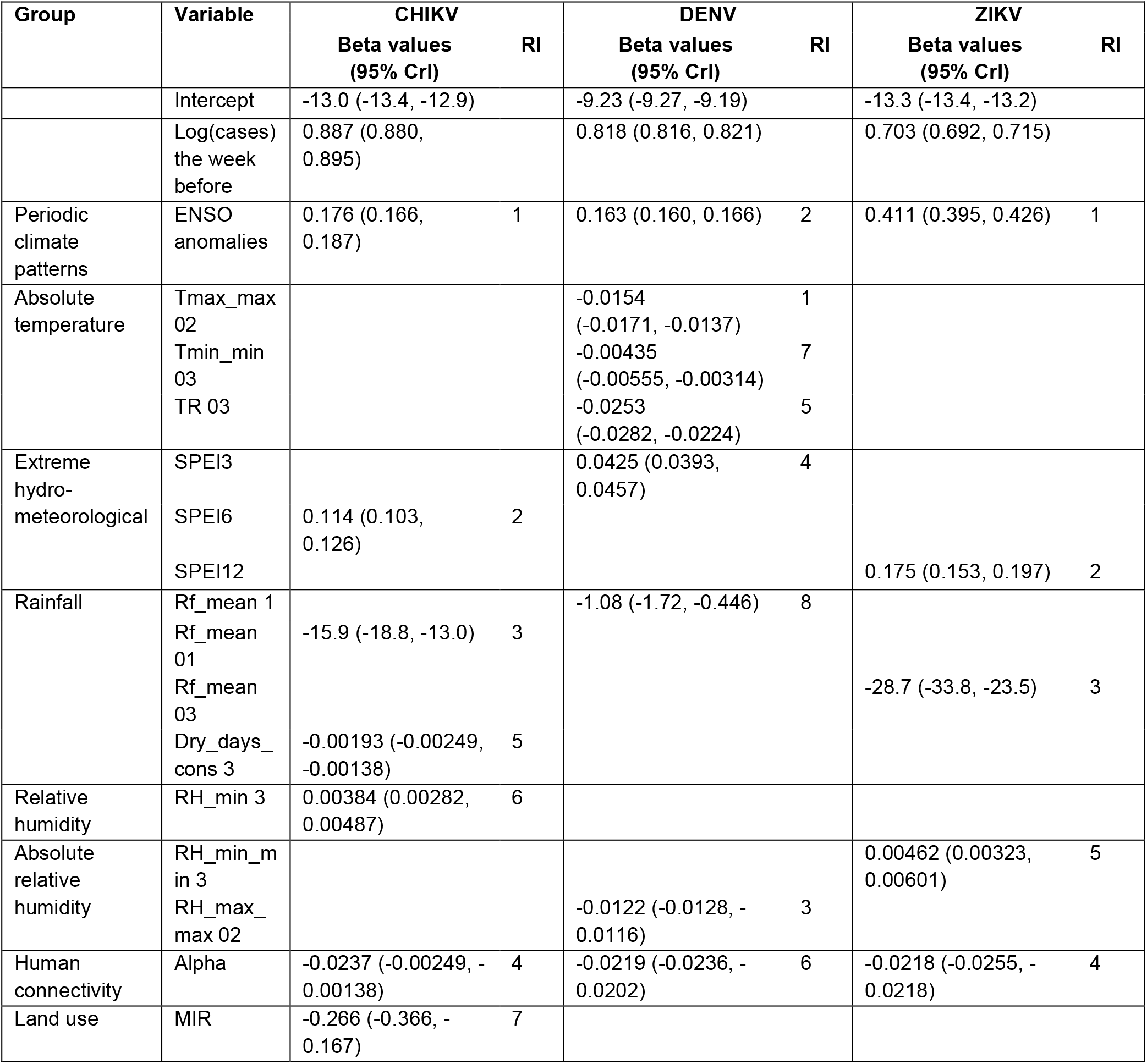
Variables in the final models. The beta coefficients and corresponding 95% Credible Intervals (CrI) are shown for the chikungunya (CHIKV), dengue (DENV), and Zika (ZIKV) models. Relative importance (RI) shows the order the variables were chosen in the model building process.

Across all models and time lags, temperature range (TR) and absolute maximum temperatures (Tmax_max) were negatively associated with arbovirus incidence (Figure 3). Tmax_max 02, Tmin_min 03, and TR 03 were predictors in the final multivariable DENV model. Tmin and Tmin_min were positively associated with CHIKV and ZIKV incidence across all time lags, however, temperature variables were not among the best predictors chosen in the final ZIKV or CHIKV models. In the univariable models, summary statistics such as Tmax_max and TR fitted better to arbovirus incidence data than Tmean (Figure 4). Environmental relative humidities, particularly minimum (RH_min), mean (RH_mean), and absolute minimum (RH_min_min), were in the top ten best predictors for CHIKV and ZIKV incidence, and were positively associated (Tables 1 and S1, Figure S1). Absolute maximum humidity (RH_max_max 02) was negatively associated with DENV incidence in the final multivariable model (Table 1).

**Figure 3.**
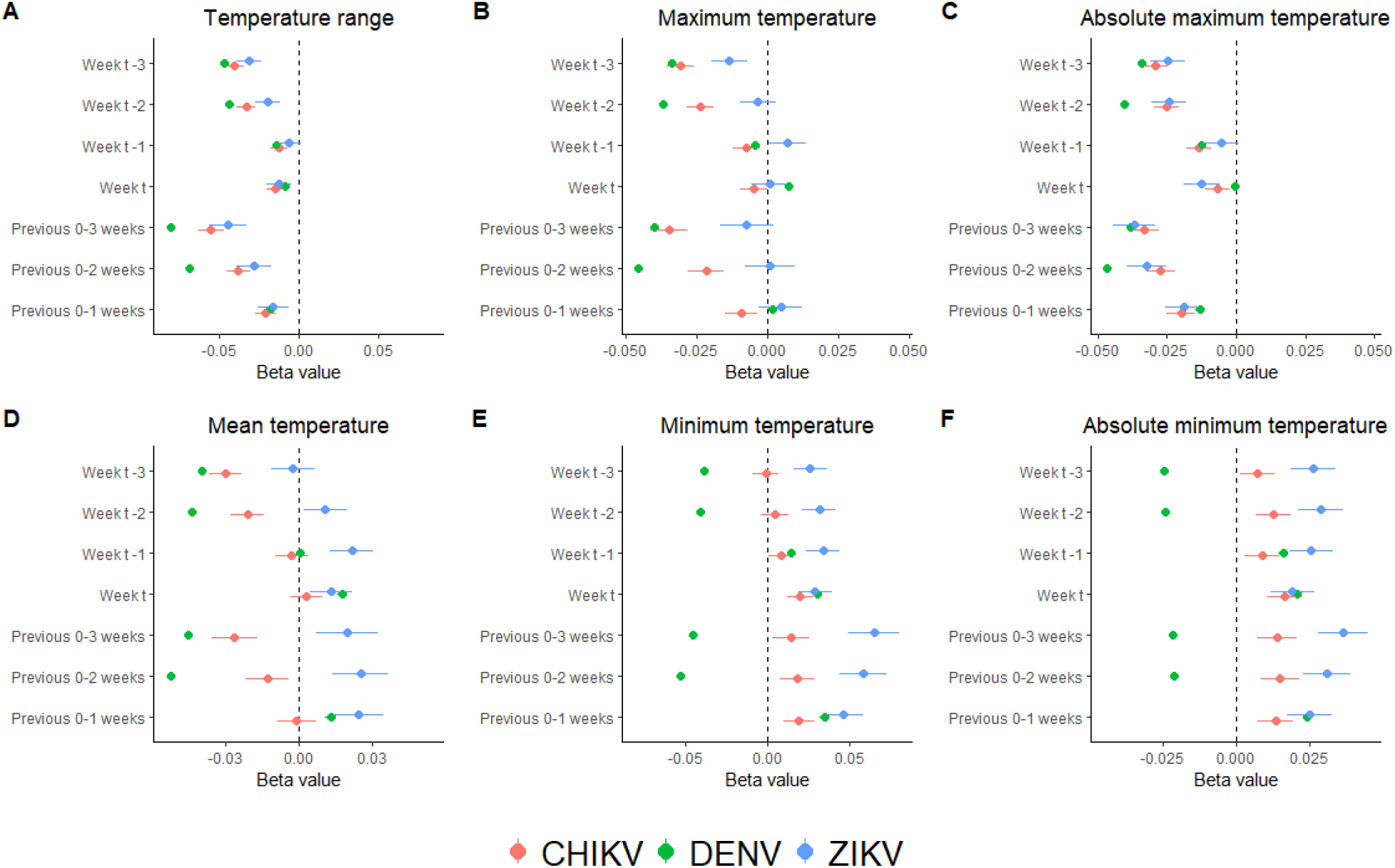
Regression coefficients for each explanatory temperature variable from the univariable models. The coefficients (beta values) are shown for the models applied to the dengue (DENV, green), chikungunya (CHIKV, red), and Zika (ZIKV, blue) data, using the average range in daily temperature (A), average maximum temperature (B), absolute maximum temperature (C), average mean temperature (D), average minimum temperature (E), absolute minimum temperature (F) across the municipalities for each week.

**Figure 4.**
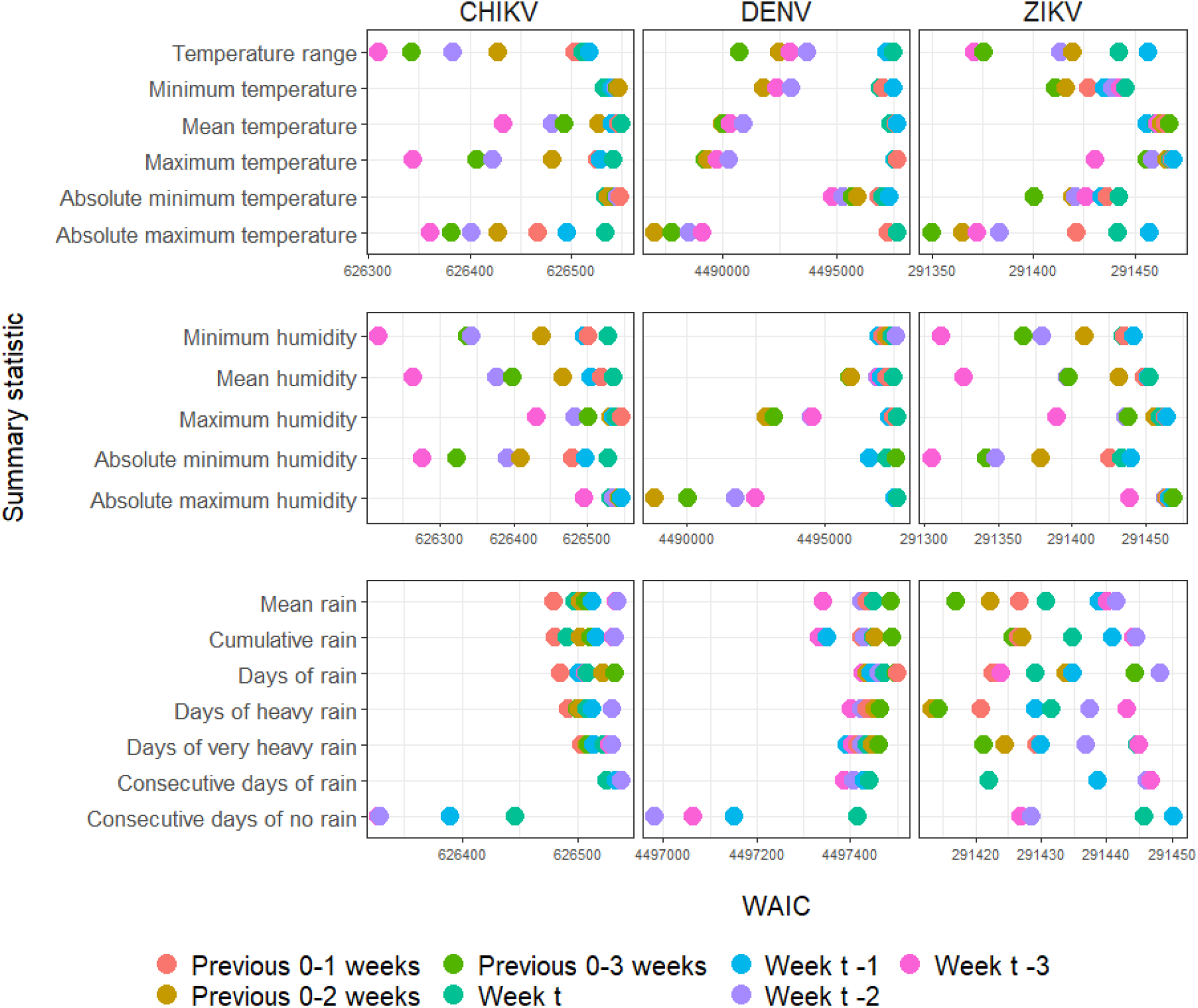
Comparison of the WAIC (Wantanabe Akaike Information Criterion) score of the chikungunya (CHIKV), dengue (DENV), and Zika (ZIKV) univariable models obtained by including each meteorological variable separately. The x-axis is ordered from lowest to highest WAIC score (best fitting to worst fitting model). The points are coloured by the time period the variable was calculated over.

Short-term summaries of mean rainfall (Rf_mean) were negatively associated with arbovirus incidence in the CHIKV, DENV, and ZIKV final multivariable models (Table 1), as was the number of consecutive days without rain (drydays_cons 3) in the CHIKV model. In the univariable models, drydays_cons fitted better to CHIKV incidence than mean and cumulative rainfall variables (Figure 4). In all the models, SPEI (which captures long-term extreme wetness or dryness) was positively associated with arbovirus incidence and had high relative importance: it was the second variable selected in the CHIKV and ZIKV final models and fourth in the DENV model (Tables 1 and S2).

Alpha centrality (which measures how connected a municipality is within the human mobility network) was negatively associated with arbovirus incidence in all the models. Tables 1 and S1-S3 compare the variables in the three final multivariable models. We observed consistent associations between arbovirus incidence and the explanatory variables across all datasets in the sensitivity analysis (Table S3).

The mean absolute error (MAE, mean of the absolute value of the estimated incidence per 100,000 persons – observed value, per municipality across the whole timeseries) was 2.90 (95% CI: 0.184, 32.1), 8.26 (95% CI: 0.996, 37.9), and 1.69 (95% CI: 0.152, 44.7) for the final CHIKV, DENV, and ZIKV models respectively. The models could capture the temporal patterns of incidence well; however peak incidence was overestimated (Figures 5 and S2-S4).

**Figure 5.**
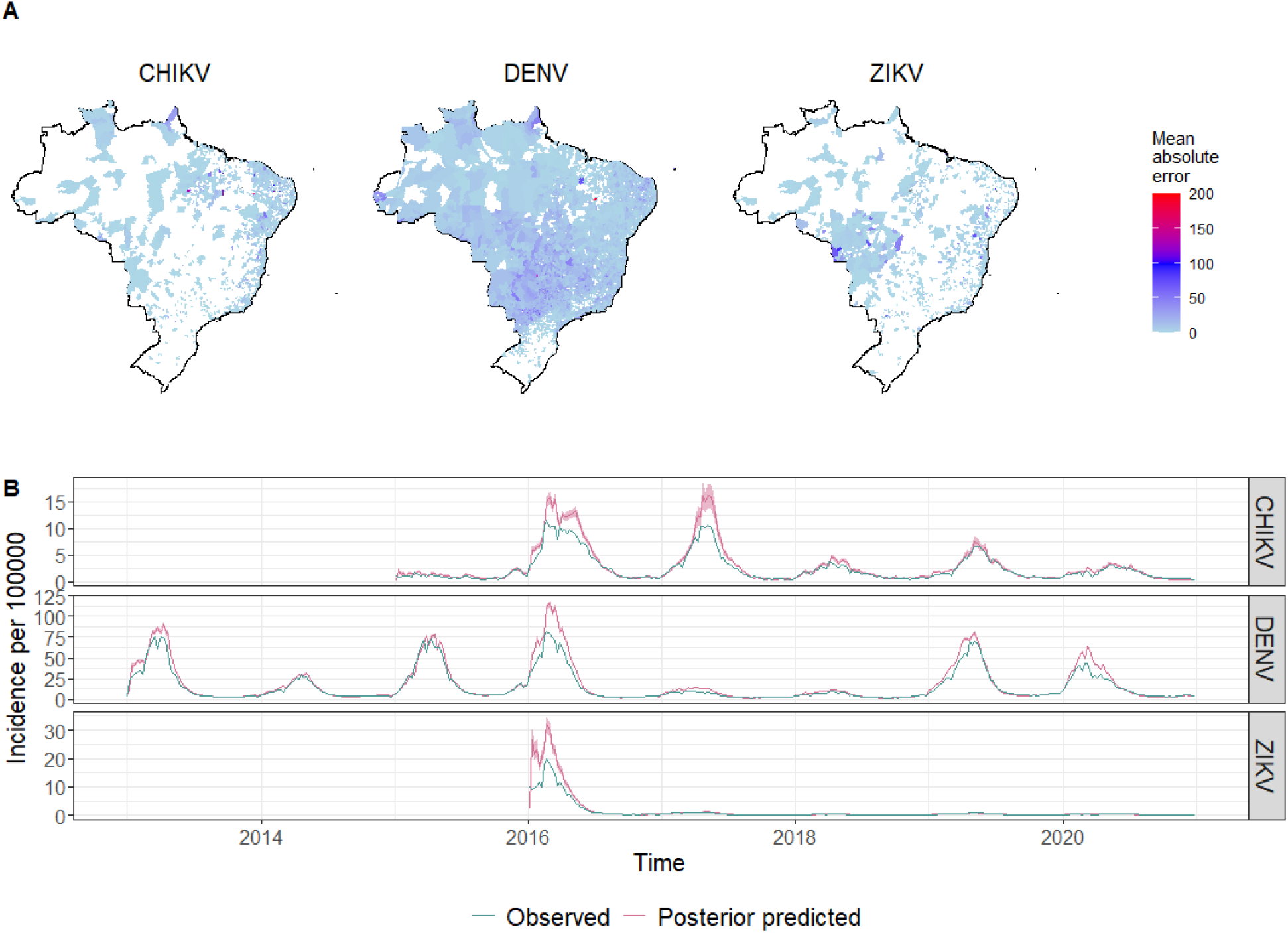
Mean absolute error and model fit for the CHIKV, DENV, and ZIKV final multivariable models. (A) Mean absolute error of model estimates for each municipality (absolute value of observed incidence per 100,000 persons – estimated incidence per 100,000 persons, summed over the weeks and divided by the number of weeks). Municipalities not included in the analysis are coloured white. (B) Comparison of the median expected incidence (red) and 95% credible intervals (CrI, in pink) compared to the observed incidence (blue) given as number of cases per 100,000 population, aggregated to the national level, obtained from the fit of the models at the municipality level.

## Discussion

We analysed weekly reported case notification timeseries data for CHIKV, DENV, and ZIKV over 2013 to 2020 at the municipality in Brazil. We explored the spatiotemporal patterns of transmission and tested the associations between incidence and variables describing meteorological, environmental, socioeconomic, human connectivity, and long-term climate conditions. We compared the associations of meteorological variables summarised over different time periods and at their average and absolute values.

The observed regularity in the timing of arbovirus epidemics across annual seasons with spatial heterogeneity (Figures 1 and 2) is consistent with previous works, for example, Churakov *et al*. 2019 showed that in Brazil between 2001 and 2016 dengue disease occurred in travelling waves each transmission season starting in the West before reaching the Northeast, with only the two most northern states not following a regular pattern ^22^ – dynamics which are also present in the 2013 to 2020 timeseries analysed in this work (Figure 2). While the timing of outbreaks, particularly of DENV, were regular between seasons, the magnitude varied with clear reductions in 2017 and 2018 (Figures 1B, 5B) as previously characterised after the 2015-2016 ZIKV epidemic ^17,32^. Immunological interactions between ZIKV and DENV have been hypothesised to be potential mediators for these patterns ^33,34^. The minimal observed ZIKV incidence after 2016 is expected to be caused by population immunity due to high levels of asymptomatic exposure during the 2015-2016 epidemic ^35^.

### ENSO anomalies and environmental temperatures

Our regression analysis results showed that global periodic climate patterns as measured by the ENSO anomalies are positively associated with arbovirus incidence across Brazil. This is consistent with previous analyses conducted in Recife during 2001 to 2017 ^36^ and Tocantins from 2010 to 2019 ^37^. The El Niño event of 2015-2016 (where positive ENSO anomalies were highest, Figure S5) has been previously linked with increased global disease outbreak risk including plague in Colorado and New Mexico, cholera in Tanzania, and DENV in Brazil and southeast Asia ^38^.

Maximum temperatures (as described by Tmax, Tmax_max, and TR variables) were negatively associated with incidence in all the univariable models (Figure 3) as well as the DENV multivariable model. Laboratory experiments consistently demonstrate that mosquito life-traits determining egg-laying, development, and feeding behaviours are non-linearly dependent on temperature, and extreme maxima limit mosquito survival ^26,39,40^. Furthermore, epidemiological studies have shown that DENV incidence can increase with temperature, but only up to a limiting maximum which was estimated at around 32 degrees Celsius in Mexico, beyond which incidence started to decline ^41^. This explains why our absolute maxima temperature variables are negatively associated; they may be capturing a threshold effect outside of optimum conditions.

### Relative humidities

We observed similar threshold patterns in the associations between arbovirus incidence and humidity as with the temperature variables: RH_min and RH_min_min variables were positively associated with incidence (in the final CHIKV and ZIKV models respectively), whilst RH_max_max was negatively associated (in the final DENV model). Relative humidity has been suggested to affect the evaporation rate and surface tension of aquatic larval habitats, which in turn impacts the probability of larval survival and therefore adult mosquito emergence. However, the mechanistic effect of humidity on mosquito hydro-regulation, fitness, and survival still needs to be fully characterised empirically ^28^.

### Rainfall patterns

Standardized Precipitation Evapotranspiration Indices (SPEI, a measure of long-term wetness or drought) were positively associated with arbovirus incidence in our models, which is consistent with Lowe *et al*. 2021 who showed that DENV risk was increased after periods of extremely wet conditions and after drought at the microregion level in Brazil between 2001 and 2019, especially in urban areas ^23^. This could be explained by increased availability of artificial mosquito breeding habitats from water storage containers in urban areas compared to rural areas during drought periods ^31^. Short-term rainfall measures such as Rf_mean were included in all three final models, where they were negatively associated with arbovirus incidence, which could be capturing the effects of decreased mosquito abundance due to flushing of rain-fed natural breeding habitats by heavy rainfall ^30^. Gibb *et al*. 2023 similarly showed that both short-term excessive rainfall and long-term drought impacted DENV risk in Vietnam, and that the risk from drought was reduced where water supply was improved ^42^. More work is needed to explore the interconnected relationships between rainfall, mosquito habitat availability, infrastructure, and human water storage behaviours at the local level over different time periods and extreme weather events. The dataset of extreme weather events that we used in this analysis recorded only the most severe events (involving for instance more than ten deaths and at least 100 people), which posed limitations to our ability to investigate the impact of serious flooding events that did not cause deaths.

### Temporal effects

Almost all the meteorological variables included in the final multivariable models were either lagged by three weeks or calculated over the previous three weeks. Furthermore, we observed a temporal effect in our univariable models, where the regression coefficients on temperature and humidity variables were further from zero when lagged by two or three weeks compared to when lagged by zero or one weeks (Figures 3 and S1). The consistent pattern in the associations suggests the variables impact different stages of arbovirus transmission cycles over the lagged time periods, for example during the larval versus adult stages. From the line-list data we calculated the average onset-to-reporting delay which was 3 days (95% CI: 0, 22 days), 3 days (95% CI: 0, 16 days), and 2 days (95% CI: 0, 19 days) for CHIKV, DENV, and ZIKV respectively. These estimates of the onset-to-reporting delay combined with an average delay between infection to symptom onset of 2, 9, and 8 days for CHIKV, DENV, and ZIKV respectively ^43–45^, and approximately 4.0, 5.5, and 6.5 days of mosquito intrinsic incubation period for CHIKV, DENV, and ZIKV ^43,44,46^ explain why lags of 2-3 weeks prior to the reported arbovirus cases can capture meteorological effects on the mosquito life-cycle stages and infection.

### Sociodemographic variables and human mobility

Our analysis suggests that human connectivity plays an important role in arbovirus transmission, as we found that alpha centrality (which measures the connectivity and relative influence of municipalities in our network analysis of human mobility data) was always negatively associated with arbovirus incidence, suggesting that less connected municipalities with smaller populations and reduced mobility do not contribute as much as larger urban centres to exporting locally acquired infections. The effect of mobility on arbovirus transmission is likely non-linear, as suggested by Lee *et al*. 2021 ^18^. Furthermore, Harish *et al*. 2024 showed that DENV invasion into the largest Brazilian city centres prior to 2020 required multiple invasions from lower population-density neighbours ^9^. Another possible explanation is that municipalities with high levels of inwards movement are usually those with better socioeconomic indicators (Figures S6-S7), and therefore the negative associations we see between arbovirus incidence and centrality could represent infrastructure and poverty-related mechanisms on transmission. This is consistent with reduced median household income, increased adult illiteracy rates, and reduced proportion of households with access to adequate sanitation being associated with increased arbovirus incidence in our univariable models. For ZIKV infection in particular, which can be transmitted sexually as well as via mosquitoes ^47^, the impact of non-meteorological variables such as demographics and human mobility likely play an important role. Other works show that increased poverty is linked to increased transmission, for example, ZIKV incidence was negatively associated with the level of education in the northeast of Brazil in 2016 ^48^ and DENV incidence was negatively associated with mean income in Cambé in southern Brazil between 2012-2014 ^49^.

### Limitations

The analysis performed in this paper was based on incidence data, which account for a portion of the true number of infections since only symptomatic and reported cases are captured by surveillance. Furthermore, dengue, Zika, and chikungunya share similar symptoms together with other circulating arboviruses not included in this analysis, such as Mayaro and Oropouche, which may have caused case misdiagnosis ^50^. Whilst under-reporting can vary in time and space due to changes in the healthcare infrastructure, the flexible spatiotemporal random effects included in our baseline models – specifically the random walk on week number (n = 53) per state (n = 27) and an adjacency smoothed random intercept on municipality (n = 1507, 4417, and 986, for CHIKV, DENV, and ZIKV) per year (n = 6, 8, 5, for CHIKV, DENV, and ZIKV) - could capture some of these differences (Figures S8-S11).

Furthermore, the temporal lag term in our models captured typical epidemic dynamics by linking generation of infections in sequential weeks. Despite this, mechanistic effects such as population immunity, cross-immunity between arboviruses, and age-structure were not explicitly accounted for in our models, and in future work it would be interesting to assess the extent to which additional information on the age of the reported cases as well as immunity (e.g. from seroprevalence surveys) affects our understanding on the impact of extreme weather conditions on the transmission dynamics of these diseases.

A limitation of the variables in this analysis is that the socioeconomic data was collated from the most recent available census which was conducted in 2010. In future work, it will be interesting to revisit the relationships between poverty and arbovirus incidence once up-to-date census estimates become available. Furthermore, we explored the impact of range and absolute temperature to represent extremes and deviations from normal meteorological patterns, in keeping with the core measures of extreme climate as selected by the Joint CCl/CLIVAR/JCOMM Expert Team on Climate Change Detection and Indices ^51^, however future work could explore alternative representations of extremes such as binary exceedance of thresholds, as well as the inclusion of non-linearity in the meteorological variables representing extremes.

### Summary

This work uses epidemiological arbovirus incidence data to investigate climate, environment, socioeconomic and mobility dependencies at fine-scale resolution across the whole of Brazil. The analysis presented in this paper shows that El Niño Southern Oscillation anomalies and extreme wetness and drought (as measured by SPEI) may drive increased arbovirus infections outside of regular seasonal patterns. This result suggests potential roles of water storage and management practices in arbovirus transmission across Brazil. Furthermore, centrality of human connectivity networks and threshold environmental temperatures and humidity are other key drivers of arbovirus infections at fine-scale temporal and spatial resolution. Notably, our results show that temperature variations and extremes are more strongly associated with arbovirus incidence than mean temperature, which is typically used as a predictor in modelling work. This brings new evidence on the health impact of extreme weather conditions on arbovirus transmission in local settings and the importance of developing models at high spatiotemporal resolution. Better understanding how arbovirus incidence is influenced by climate, meteorology, environment, and socioeconomic conditions as well as demography at the local level is a key priority which has important implications for both public health and climate policy and planning.

## Materials and methods

### Data sharing

The individual line-list data were obtained from the Brazilian Ministry of Health via the national information system for notifiable diseases - http://portalsinan.saude.gov.br (SINAN: Sistema de Informação de Agravos de Notificação), which is publicly available upon request.

Aggregated, non-identifiable case notification data at the weekly and municipality level, along with the corresponding collated variables from our analysis are made available at the following Zenodo repository: 10.5281/zenodo.13286671. Code to run the analyses are made available at the following GitHub repository: https://github.com/mrc-ide/Brazil-arbovirus-meteo.

### Data

#### Case notification data

Daily reported cases of CHIKV, ZIKV and DENV provided by the Ministry of Health of Brazil between 2015-2020, 2016-2020 and 2013-2020, respectively (Figure 1), were acquired through the national information system for notifiable diseases (SINAN: Sistema de Informação de Agravos de Notificação). The data were aggregated to weekly time-steps at the municipality level, where the municipality recorded was the municipality of suspected infection for 64%, 58%, and 66% of the data entries in the CHIKV, DENV and ZIKV datasets respectively which reported this information, and for the remaining data entries we used the municipality of residence. We imputed zeroes for the days in between the first and last date of reported cases for each municipality. To capture local transmission and exclude the municipalities with imported cases but no active transmission, we only included municipalities which reported >=5 cases in each year or >20 cases in any one individual year. In sensitivity analyses we tested the effect of aggregating the data using (i) the municipality of residence for all data entries, (ii) the municipality where a case was reported or hospitalised for all data entries, or (iii) the municipality of suspected infection where available and the municipality of residence otherwise, as in the main analysis, except without filtering municipalities.

#### Population data

Annual global population data was extracted from LandScan™ datasets at approximately 1/120 degrees resolution for 2013 to 2019 ^53–60^. The 2020 data were unavailable, and we assumed 2020 values were the same as 2019. The pixel level data were extracted and aggregated over the Brazilian municipalities ^61^.

#### Human connectivity data

The mobility data were collected by a private Brazilian company (Incognia - https://www.incognia.com/pt/politicas/covid) that gathered anonymous geolocation of users of mobile apps that used the company’s software development kit. The data were available through a research agreement. Original anonymous data was provided as described in Peixoto *et al*., 2020 ^62^, considering journeys as origin-destination latitude-longitude pairs, which were aggregated at the municipality and monthly timescale to estimate three measures of centrality; degree, betweenness, and alpha centrality (see Supplementary Methods).

#### Meteorological data

Global hourly data for total precipitation, 2m dew point temperature, and 2m temperature variables between 1 January 2013 to 31 December 2020 were downloaded from the ERA5 reanalysis dataset provided by Copernicus Climate Change Service at 0.1 × 0.1 degrees resolution ^63^. Relative humidity (RH) was calculated using the dew point and temperature (Supplementary Methods). The pixel level meteorological data were population weighted to better capture the climate and environmental conditions that the average person in each municipality experiences, which discounts the conditions where there is no population and weights populous locations more compared to less populated areas (Supplementary Methods).

#### ENSO data

El Niño Southern Oscillation (ENSO) is a periodic cycle in sea surface temperatures (SST) in the Pacific Ocean which determines whether El Niño or La Niña events are occurring and impacts the atmosphere and global weather conditions. Monthly SST anomalies in the east-central equatorial Pacific region 3.4 (Niño 3.4, 5N-5S, 170-120W), which are defined as departures from the long-term average monthly temperature between 1991 to 2020, were obtained from the National Oceanic and Atmospheric Administration (psl.noaa.gov/data/correlation/nina34.anom.data) ^64^ for January 2013 to December 2020.

#### Extreme weather events data

Disaster events for Brazil between 2013 and 2020 were obtained from the International Disaster Database ^65^. Disaster events are classified as those where 10+ people were killed, 100+ people are affected, there is a call for international assistance or a declaration of a state of emergency. Disaster events which led to flooding (e.g., storms, landslides, and the dam breaks in Minas Gerais in 2015 and 2019) were recorded. The raw disaster data was given at either the municipality level or the state level (in which case we assumed that every municipality in the state experienced the disaster event). Categorical variables for each week in 2013 to 2020 were created which described if there had been a flooding event during the previous 1, 2, and 3 weeks.

#### Socioeconomic data

Data for the median household income, illiteracy rate (percentage of people over 15 years old who cannot read or write), and sanitation (percentage of people in houses with adequate sanitation) per municipality, were collated from the most recent published census conducted in 2010 ^66^.

#### Land-use data

Mid-infrared (MIR) data collected every eight days between 2013 and 2020, was sourced from NASA’s Moderate Resolution Imaging Spectroradiometer (MODIS) satellite instruments MOD13A2v061 and MYD13A2v061 ^67,68^. Vegetated surfaces, agricultural land and urban settlements differentially reflect MIR. The MIR data was scaled to give a value between 0 and 1. The MIR data was population-weighted and averaged for each municipality of Brazil per week (Supplementary Methods).

### Calculating meteorological indices

We aggregated the hourly data to calculate population-weighted weekly average mean, maximum, and minimum relative humidity (RH_mean, RH_max, RH_min), average mean, maximum, and minimum temperature (Tmean, Tmax, Tmin), absolute maximum of maximum relative humidity (RH_max_max), absolute minimum of minimum relative humidity (RH_min_min), absolute maximum of maximum temperature* (Tmax_max), absolute minimum of minimum temperature* (Tmin_min), and average temperature range* (TR). We also calculated the number of rain days where daily cumulative precipitation exceeded 1mm (raindays), the number of heavy rain days (precipitation >10mm)* (raindays_heavy), the number of very heavy rain days (precipitation >20mm)* (raindays_heavy+), average precipitation (mean of rain on days where precipitation >1mm) (Rf_mean), cumulative precipitation (Rf_cum), and the maximum number of preceding consecutive rain days (precipitation >1mm)* and dry days (precipitation <1mm)* (raindays_cons and drydays_cons) across the timeseries. Variables denoted with an asterisk (*) are core measures of extreme climate as selected by the Joint CCl/CLIVAR/JCOMM Expert Team on Climate Change Detection and Indices ^51^. For each week t, the variables were calculated at time lags of 0 (t), 1 (t-1), 2 (t-2) and 3 (t-3) weeks, and over the cumulative time periods t to t-1, t to t-2 and t to t-3.

The meteorological data was aggregated to the monthly level and used to calculate the Standardized Precipitation Evapotranspiration Index (SPEI), a measure of long-term wetness or dryness/drought across different time-scales (1-, 3-, 6- and 12-months) using the *SPEI* R package ^69^ (Supplementary Methods).

### Spatiotemporal Bayesian mixed effects regression modelling

We developed hierarchical spatiotemporal Bayesian mixed-effects models for the number of reported cases of CHIKV, ZIKV, or DENV in municipality *i* at week *t* (Y_*i, t*_). We started by developing ‘baseline’ models capturing seasonal and spatial variation in transmission, and then developed ‘univariable’ models including a single explanatory variable, followed by ‘multivariable’ models including more than one explanatory variable. We assumed that the reported number of cases (*Y*_*i, t*_) followed a negative binomial distribution (Equation 1), where *κ* is the overdispersion parameter, *Z*_*i, t*_ is the population in municipality *i* at week *t* (included as an offset in the models) and *P*_*i, t*_ is the linear predictor.

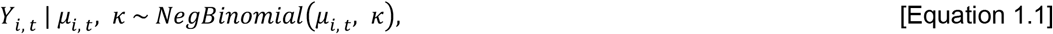

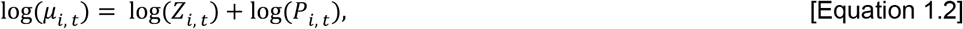

The linear predictor in the baseline models was modelled as shown in Equation 2, where *β*_0_ represents the intercept, *β*_1_*Y*_*i, t*−1_ is a lag term specifying the number of reported cases in municipality *i* the week before 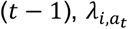 is a spatial random effect on the municipality *i* per year *a*_*t*_, and 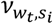 is a temporal random effect on the week number (*w*) per state (*s*) of the municipality it belongs to.

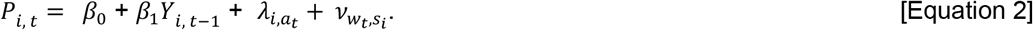

The random effect on the week number 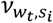 captures regular seasonality (intra-annual dynamics) and was included as a cyclic random walk of order 1 (Figure S8). The random effect on municipality 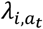 captures spatial correlation and long-term trends over time (inter-annual dynamics), for example differences in surveillance capacity between locations, which was modelled using a modified Besag-York-Mollié model (bym2) that accounts for unstructured (independent noise) as well as structured (spatial dependencies) effects, by smoothing the variables according to their defined neighborhood/adjacency structure (Figures S9-11). The neighbourhood matrices were generated from the administrative level 2 shapefile of Brazil in the *geobr* package using the *spdep* package ^70,71^.

The regression model parameters were estimated using the *R-INLA* package ^72,73^, which uses Integrated Nested Laplace Approximation for model calibration. The results from this analysis were visualised using the R packages *ggplot2, geofacet*, and *sf* ^74–76^.

### Initial variable selection

The linear predictor in the baseline models was modified to include one extra variable (A) to generate univariable models (Equation 3, where *X*_*i,t*_ is a continuous explanatory variable with coefficient *β*_2_, and *ψ*_*i, t*_represents a categorical explanatory variable, with a coefficient *β*_2:*n*_ per category, *n*) separately for each of the meteorological, land use, socioeconomic, centrality, extreme weather, and ENSO explanatory variables (n = 139), which are described and visualised in the Supplementary Materials (Table S4, Figures S5-S7, S12-S17).

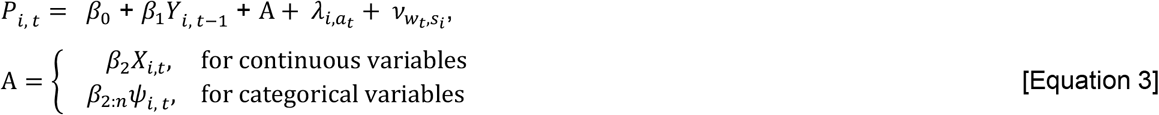

We determined the significance of the association of the explanatory variable with the response variable through the 95% Credible Intervals (CrI) of the regression coefficient which was considered not significant when it crossed zero. Variables that were not significantly associated in this initial selection step were not used in further model development for the multivariable models described later.

For each univariable model, we extracted the Watanabe-Akaike Information Criterion (WAIC) value which is an estimate of the prediction error ^77^. For the lagged meteorological variables, only the lag whose corresponding univariable model had the lowest WAIC score (the best-fitting) was used in later model development (see flow diagram in Figure S18), for example if the best fitting minimum temperature DENV univariable model used lag of week t-2, then minimum temperature at t, t-1, t-3, t to t-1, t to t-2, and t to 1-3 were not used in development of DENV multivariable models.

### Multivariable model development

The explanatory variables selected during the univariable analysis were used to build multivariable models in a stepwise forward selection approach (Figure S18). The multivariable linear predictors were modelled as shown in Equation 4, where *X*_*j,i,t*_ are continuous explanatory variables with *j* = 1, …, *n*, denoting the number of variables included each with coefficient *β*_*j*+1_. Only continuous explanatory variables were considered, because no categorical variables were retained after the univariable analysis.

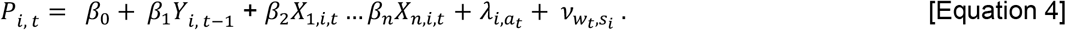

The multivariable models were built in steps (Table S2) where at each step, variables were added one at a time to the models and included if: (1) they improved the WAIC score more than the other variables tested in that step, (2) the WAIC improvement compared to the previous step was >4, (3) the variable was not collinear with any variables already included in the model from previous steps (defined as having an absolute Pearson’s correlation coefficient >0.6), and (4) the variable was significantly associated with the response variable (the 95% CrI of the *β* coefficient did not cross zero).

In a sensitivity analysis we fitted the final multivariable models to incidence data where the municipality of a case was assigned using the municipality of residence or the municipality where a case was reported or hospitalised (Table S3).

### Assessment of model performance

We computed the mean absolute error (MAE, defined as the absolute value of the median estimated incidence minus the observed incidence, averaged across the whole time series) for each municipality. We summarised the MAE nationally by presenting the 0.50, 0.025, and 0.975 quantiles of the municipality level MAE. To visualize the model fit, we sampled 100 realisations from the posterior distribution of the model, estimated the incidence at the state and national level, and extracted the 0.50, 0.025, and 0.975 quantiles to generate the median and associated 95% CrIs.

## Data Availability

The individual line-list data were obtained from the Brazilian Ministry of Health via the national information system for notifiable diseases - http://portalsinan.saude.gov.br (SINAN: Sistema de Informação de Agravos de Notificação), which is publicly available upon request. Aggregated, non-identifiable case notification data at the weekly and municipality level, along with the corresponding collated variables from our analysis are made available at the following Zenodo repository: 10.5281/zenodo.13286671. Code to run the analyses are made available at the following GitHub repository: https://github.com/mrc-ide/Brazil-arbovirus-meteo.

https://doi.org/10.5281/zenodo.13286670

## Acknowledgements

We acknowledge the advice and input from Dr Sally Jahn in interpreting the meteorological data. We acknowledge the use of meteorological data from Muñoz Sabater, J. (2019), which was downloaded from the Copernicus Climate Change Service (C3S) Climate Data Store: ERA5-Land hourly data from 1950 to present. The results contain modified Copernicus Climate Change Service information 2023. Neither the European Commission nor ECMWF is responsible for any use that may be made of the Copernicus information or data it contains.

